# A Non-Adaptive Combinatorial Group Testing Strategy to Facilitate Healthcare Worker Screening During the Severe Acute Respiratory Syndrome Coronavirus-2 (SARS-CoV-2) Outbreak

**DOI:** 10.1101/2020.07.21.20157677

**Authors:** JH McDermott, D Stoddard, PJ Woolf, JM Ellingford, D Gokhale, A Taylor, LAM Demain, WG Newman, G Black

**Affiliations:** Manchester Centre for Genomic Medicine, St Mary’s Hospital, Manchester University NHS Foundation Trust, Manchester, M13 9WL, UK; Division of Evolution and Genomic Sciences, School of Biological Sciences, University of Manchester, Manchester, UK; DS Analytics and Machine Learning Ltd, 12 Hammersmith Grove, Hammersmith, London, M6 7AP; Origami Assays, Ann Arbor, Michigan, USA

## Abstract

**Background:** Regular SARS-CoV-2 testing of healthcare workers (HCWs) has been proposed to prevent healthcare facilities becoming persistent reservoirs of infectivity. Using monoplex testing, widespread screening would be prohibitively expensive, and throughput may not meet demand. We propose a non-adaptive combinatorial (NAC) group-testing strategy to increase throughput and facilitate rapid turnaround via a single round of testing.

**Methods:** NAC matrices were constructed for sample sizes of 700, 350 and 250 with replicates of 2, 4 and 5, respectively. Matrix performance was tested by simulation under different SARS-CoV-2 prevalence scenarios of 0.1-10%, with each simulation ran for 10,000 iterations. Outcomes included the proportions of re-tests required and the proportion of true negatives identified. NAC matrices were compared to Dorfman Sequential (DS) approaches. A web application (www.samplepooling.com) was designed to decode results.

**Findings:** NAC matrices performed well at low prevalence levels with an average number of 585 tests saved per assay in the n=700 matrix at a 1% prevalence. As prevalence increased, matrix performance deteriorated with n=250 most tolerant. In simulations of low to medium (0.1%-3%) prevalence levels all NAC matrices were superior, as measured by fewer repeated tests required, to the DS approaches. At very high prevalence levels (10%) the DS matrix was marginally superior, however both group testing approaches performed poorly at high prevalence levels.

**Interpretation:** This testing strategy maximises the proportion of samples resolved after a single round of testing, allowing prompt return of results to staff members. Using the methodology described here, laboratories can adapt their testing scheme based on required throughput and the current population prevalence, facilitating a data-driven testing strategy.

**Funding:** None to Declare

## 1. Introduction

Throughout the severe acute respiratory syndrome coronavirus-2 (SARS-CoV-2) outbreak there have been calls for widespread testing to help track and suppress viral transmission.^1; 2^ Many countries have adopted high-throughput testing strategies and tens of millions of SARS-CoV-2 antigen tests have been performed since the outbreak began. The reagents required to perform these tests, because of the unparalleled global demand, are a limited resource and their utilization should be optimized.

In many regions the prevalence of the SARS-COV-2-associated respiratory disease (COVID-19) is beginning to fall in the general population.^3^ However, certain settings such as hospitals and care homes, have the potential to act as persistent reservoirs of infection where the reproduction (R) value remains persistently elevated. Approaches to ameliorate nosocomial spread include access to adequate personal protective equipment, effective cohorting of patients and the proactive identification of infectious staff members.^1;4^

Where staff develop symptoms, they should isolate. However, a yet undetermined proportion of patients infected with SARS-CoV-2 develop an asymptomatic viremia.^5; 6^ These asymptomatic carriers pose a serious challenge when attempting to prevent spread within hospitals, environments where staff often congregate in close proximity with vulnerable patients. Because of this, there is a growing demand for the routine testing healthcare workers (HCWs), a premise supported by modelling which suggested screening, irrespective of symptoms, could reduce transmission by 25- 33%.^1; 7^

One of the main hurdles to initiating a comprehensive hospital staff testing program is the large number of staff requiring testing and the rapid turnaround times which would be required to make any screening strategy useful. An estimated 1.5 million staff work in the NHS, with larger hospitals each employing over 20,000 employees. The requirement to test even a small proportion of these patients would dwarf the U.K.’s testing capacity and would be prohibitively expensive using standard monoplex testing, even taking into account the estimated £5 Billion the UK government plans to spend on SARS-CoV-2 over the next 2 years.^8^ However, in moving from an individual diagnostic approach towards the population-based screening of asymptomatic individuals, we argue that there is a key shift in the philosophy underpinning the application of testing and alternative diagnostic approaches could be used, specifically a group testing strategy.

A group testing strategy is where samples taken from more than one person are tested at the same time. The principle being that if a pool returns negative, then everyone in that pool is negative. If a pool returns positive, then at least one sample in that pool is positive (Figure 1). The concept was first introduced by Dorfman in 1943, who proposed it as an approach to screen soldiers for syphilis during World War 2.^9^ Most group testing approaches used since have been based on Dorfman’s original methodology. This involves the pooling of multiple samples and, if a pool returns positive, the constituent samples undergo further testing (Figure 1).^10^ This approach will be referred to as Dorfman Sequential (DS) pooling henceforth.

**Figure 1.**
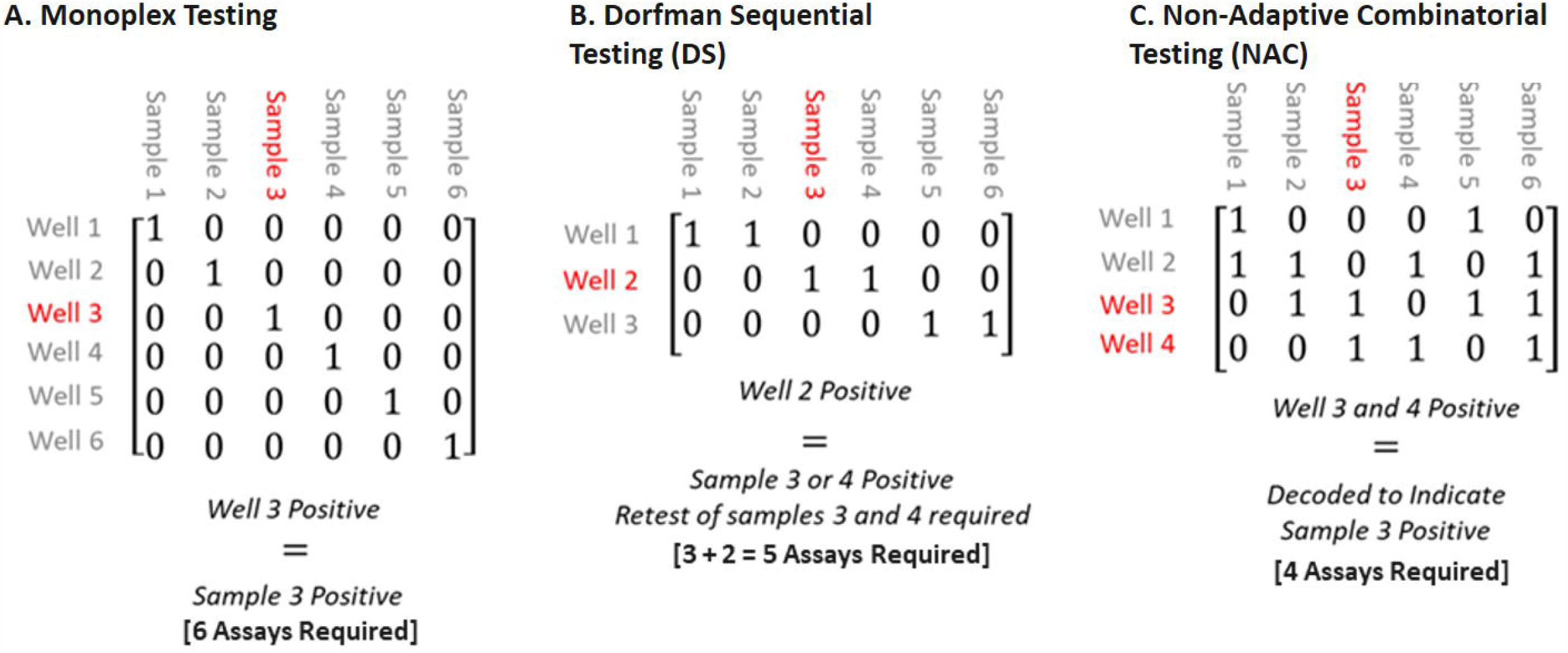
SARS-CoV-2 Testing Approaches. Three approaches are outlined with 6 samples where sample 3 is positive. (A) Where there is no pooling (monoplex testing) each well contains one sample, therefore the result from that well corresponds directly to that sample. (B) Using Dorfman Sequential (DS) pooling, wells contain more than one sample. In this example well 2 is positive which, according to the matrix, contains samples 3 and 4. Retesting of samples 3 and 4 is required to determine that sample 3 is positive (C) Using non-adaptive combinatorial strategies, sample 3 is in both wells 3 and 4 which appear positive. Given sample 3 is the only sample in both wells 3 and 4, the matrix can be decoded to indicate that sample 3 is positive without a requirement for retesting.

Although DS pooling can increase capacity significantly compared to a monoplex approach, throughput may not be maximised as at least two rounds of testing are required to differentiate positive samples within a pool. In the case of SARS-CoV-2, if the samples were pooled prior to viral RNA extraction, then a further RNA extraction step would be required for the second round of testing, significantly slowing the process. In the context of HCW screening, any lengthening of the testing process could mean a reduced workforce while staff await their results, impacting patient care.

Non-adaptive approaches (Figure 1) overcome the requirement for repeated testing rounds by testing the same sample in several simultaneously assayed pools, aiming to maximize the proportion of samples resolved after a single round of testing. Although DS approaches may ultimately require fewer tests over several rounds compared to non-adaptive approaches, in the context of HCW screening, maximizing the number of true negatives identified after a single round of testing should be the priority for any testing scheme. We contend that multi-stage group testing approaches are not most suitable for HCW screening as they do not recognize the need for rapid results. More than one round of group testing would introduce a complexity into assay design and results may be considerably delayed for some individuals if their samples are not resolved in the first round.

Herein, we suggest that a group testing approach should be used as an initial screen, maximizing the number of true negatives identified, before monoplex testing is used to determine any indeterminate samples (Figure 2). For this, we propose a non-adaptive combinatorial (NAC) pooling approach as an alternative screening strategy to maximise throughput after a single round of testing in the context of varying population prevalences of SARS-CoV-2 infection.

**Figure 2.**
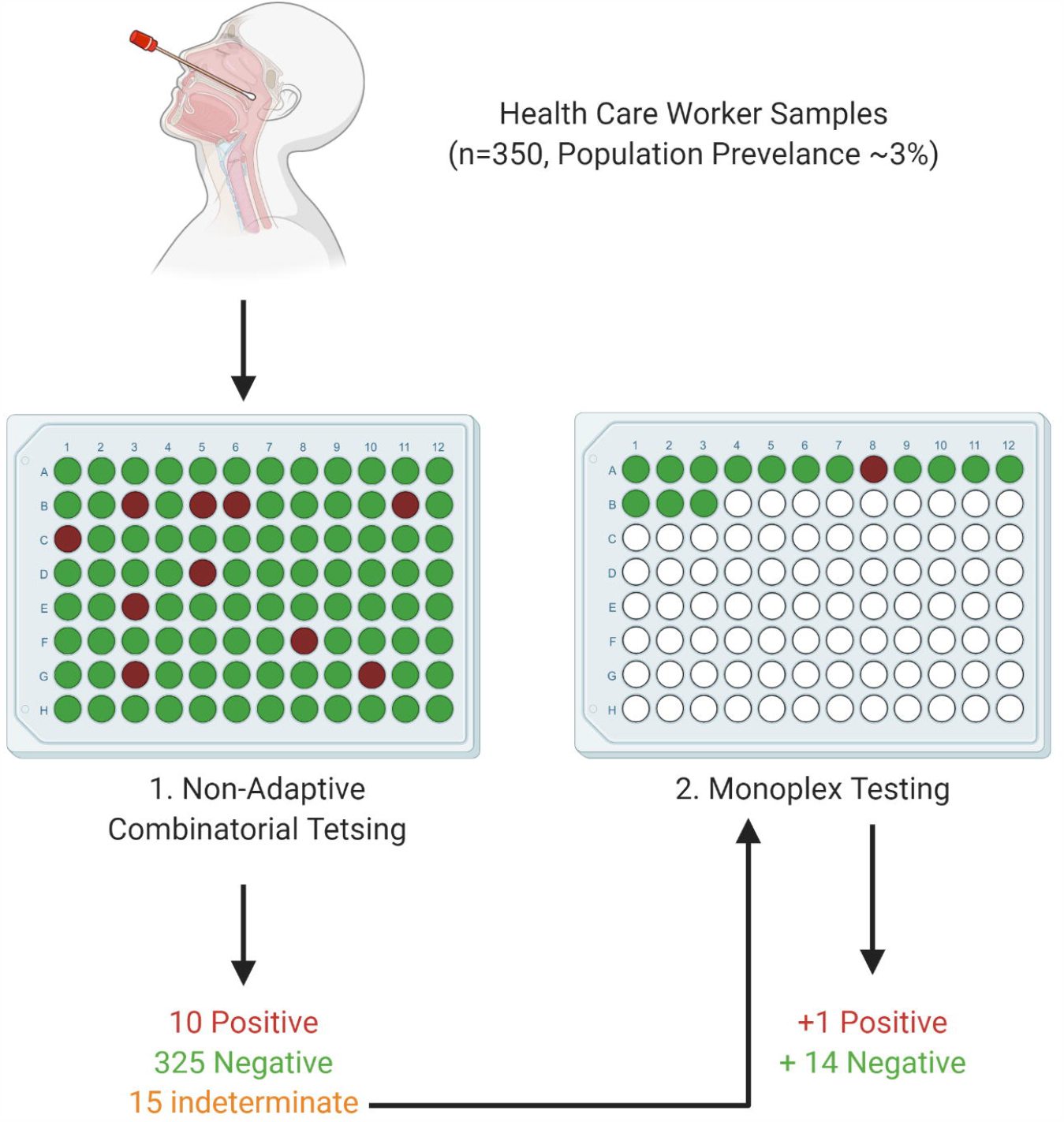
Proposed Testing Approach for Healthcare Worker Screening. A single round of group testing via a non-adaptive combinatorial scheme is used as an initial high-throughput screen. In this example, this screen can confirm the status of 335 samples (96%). The 15 indeterminate samples are then determined via monoplex testing, rather than a second group testing round being implemented.

## 2. Methodology

### 2.1 Sample Pooling to Establish Limit of Detection

To establish an approximate suitable limit of detection (LOD) for pooling, nasopharyngeal samples of known SARS-CoV-2 status were pooled prior to extraction. Two pools (Pool-1, Pool-2) were prepared each comprising 14 SARS-CoV-2 negative and one SARS-CoV-2 positive sample. Pool-1 contained a positive sample with a viral load near the limit of detection at previous testing using the IDT N-gene assay (Ct= 37). Pool 2 contained a positive sample with a mid-level viral load (Ct= 30, classified as mid-range from >100 positive clinical samples). The IDT N-Gene EUA assay comprises two targets located in the CV19 N-gene (N1 and N2). The assay has been designed and batch verified by the CDC and is run according to CDC protocol.^11^ Pooled samples were extracted using the AmoyDx Virus/Cell RNA kit (AmoyDx, Xiamen, China). RNA was tested for SARS-CoV-2 using the EUA IDT N-gene assay (IDT, CA, USA) and run on the ABI QuantStudio6 instrument (Thermofisher, MA, USA) 200 copies of 2019-nCoV_N_Positive (IDT, CA, USA) were run as a positive control. Baselines and thresholds were defined automatically by the ABI QuantStudio6 software. SARS-CoV-2 positive or negative status was assigned using the criteria defined by the CDC (Supplementary Data: Table S1).

### 2.2 Design of NAC and DS Pooling Matrices

Simulations were run using the R software package to construct NAC pooling matrices for different pre-determined input values of n, the number of samples, and w the number of wells to which each sample is allocated. The number of wells is assumed to be fixed at 96 throughout and the maximum pool size was fixed at 15, informed by the LOD study as described above. Example pooling matrices were generated for n values of 700, 350 and 250 with w values of 2, 4 and 5, respectively. The code for the matrix design algorithm is publicly accessible (https://github.com/duncstod/grouptesting).

Each pooling matrix was constructed by randomly allocating samples to wells until the following conditions are satisfied: 1) no sample is tested in the same well more than once, 2) and no sample pairs are tested together in the same wells more than once. The maximum number of guaranteed positives a pooling matrix can identify is r-1. Such matrices are r-1 disjunct. Pooling matrices can be accessed at www.samplepooling.com.

The performance of each non-adaptive matrix was tested against a matrix with a w value of 1 at the same sample size. This w=1 matrix represents the first stage of a DS testing scheme and will be referred to as the DS matrix henceforth. Pool size for the DS matricies was determined by the number of samples divided by the number of wells, and each sample was allocated to a single well.

### 2.3 Iterative Testing of NAC and DS Pooling Matrices

The efficacy of each matrix was tested by simulation under different SARS-CoV-2 prevalence scenarios of 0.1%, 1%, 3%, 7%, and 10% (Table 1). Each simulation ran for 10,000 iterations. In each iteration every sample is designated as positive or negative by n draws from a Bernoulli distribution with p = 0.1%, 1%, 3%, 7% and 10%.

**Table 1.** An outline of the 3 different non-adaptive combinatorial matrices tested at 5 different population prevalence levels. n=sample size, w= number of times the sample is repeated over the assay plate, p= population prevalence. Figures are presented as mean values from 10,000 simulations.

| Simulation Number | n | w | p | Tests confirmed negative (n) | True Negatives Confirmed (%) | Expected Retests Required (n) | Expected Tests Saved (n) |
| --- | --- | --- | --- | --- | --- | --- | --- |
| 1 | 700 | 2 | 0.1% | 699 | 99% | 1 | 603 |
| 2 |  |  | 1% | 681 | 97% | 18 | 586 |
| 3 |  |  | 3% | 601 | 86% | 100 | 504 |
| 4 |  |  | 7% | 393 | 56% | 305 | 299 |
| 5 |  |  | 10% | 264 | 38% | 435 | 169 |
| 6 | 350 | 4 | 0.1% | 350 | 100% | 0 | 254 |
| 7 |  |  | 1% | 346 | 99% | 4 | 250 |
| 8 |  |  | 3% | 335 | 96% | 15 | 239 |
| 9 |  |  | 7% | 275 | 79% | 75 | 179 |
| 10 |  |  | 10% | 209 | 60% | 141 | 113 |
| 11 | 250 | 5 | 0.1% | 250 | 100% | 0 | 154 |
| 12 |  |  | 1% | 247 | 99% | 3 | 151 |
| 13 |  |  | 3% | 242 | 97% | 8 | 146 |
| 14 |  |  | 7% | 217 | 87% | 33 | 121 |
| 15 |  |  | 10% | 182 | 72% | 69 | 85 |

From each simulation we derived pooling matrix performance statistics, tested with zero error, including the number of tests saved in comparison to a monoplex approach, measured as [*n - (indeterminate results* - 96)] where n represents the total samples tested on a 96-well plate and indeterminate results are those samples that cannot be decoded through the matrix, meaning retesting is required. The average proportion of samples confirmed as negative for each matrix was also determined.

### 2.4 Development of an Application to Decode Matrix Results

A web platform was designed (www.samplepooling.com) which allows users to choose their matrix (n=700, n=350, n=250) and decode their results. A Combinatorial Orthogonal Matching Pursuit (COMP) algorithm initially assumes that each sample is positive at the beginning of the decoding process.^12^ Attempts are then made to disprove this assertion by finding a well in which the sample has been placed which has called as negative. A Definite Defective (DD) algorithm was then used to find positive wells which contain a single sample on the list of potentially positive samples. This attributes a status for each result as either “Positive” or “Indeterminate Result” where re-analysis is suggested. If the decoding system identifies a well which is not consistent with the matrix, such as a false positive result, the results for the concordant wells are displayed but an error message is shown for the discrepant well and reanalysis is suggested. The code is publicly accessible via https://github.com/MCGM-Covid-19/matrix-decoder.github.io.

## 3. Results

In order to establish a suitable limit of detection (LOD) for pooling, nasopharyngeal samples of known SARS-CoV-2 status were placed in two pools, each comprising 14 SARS-CoV-2 negative and one SARS-CoV-2 positive sample, the positive samples at differing viral loads. Both pooled samples tested positive for SARS-Cov-2 under the Centers for Disease Control and Prevention (CDC) defined guidelines; this indicated that positive samples can be detected when diluted 15-fold (Supplementary Data). This limit was used as an assumption to inform the design of the pooling matrices.

The performance of all the DS matrices, as defined by the expected number of retests required, deteriorated as the population prevalence increased (Table 1). Matrices which were designed to test more samples were less tolerant to increases in population prevalence than those designed to test fewer (Figure 3). In the simulation where there was a population prevalence of 1%, the n=700 NAC matrix performed well, with an average of only 19 samples requiring retesting, representing an average of 585 tests saved per 96 well plate [700 - *(indeterminate results*[19] - 96) = 585]. However, as the population prevalence increased the average number of tests decreased markedly with an average of only 168 tests saved and 436 retests required when the population prevalence was at 10%. Given the matrices were tested using draws from a Bernoulli distribution, there were some simulations where the actual sample positivity rate was greater than 10%. Here the performance of the n=700 matrix deteriorated significantly, with some simulations saving almost no tests compared to no pooling approach (Figure 3).

**Figure 3.**
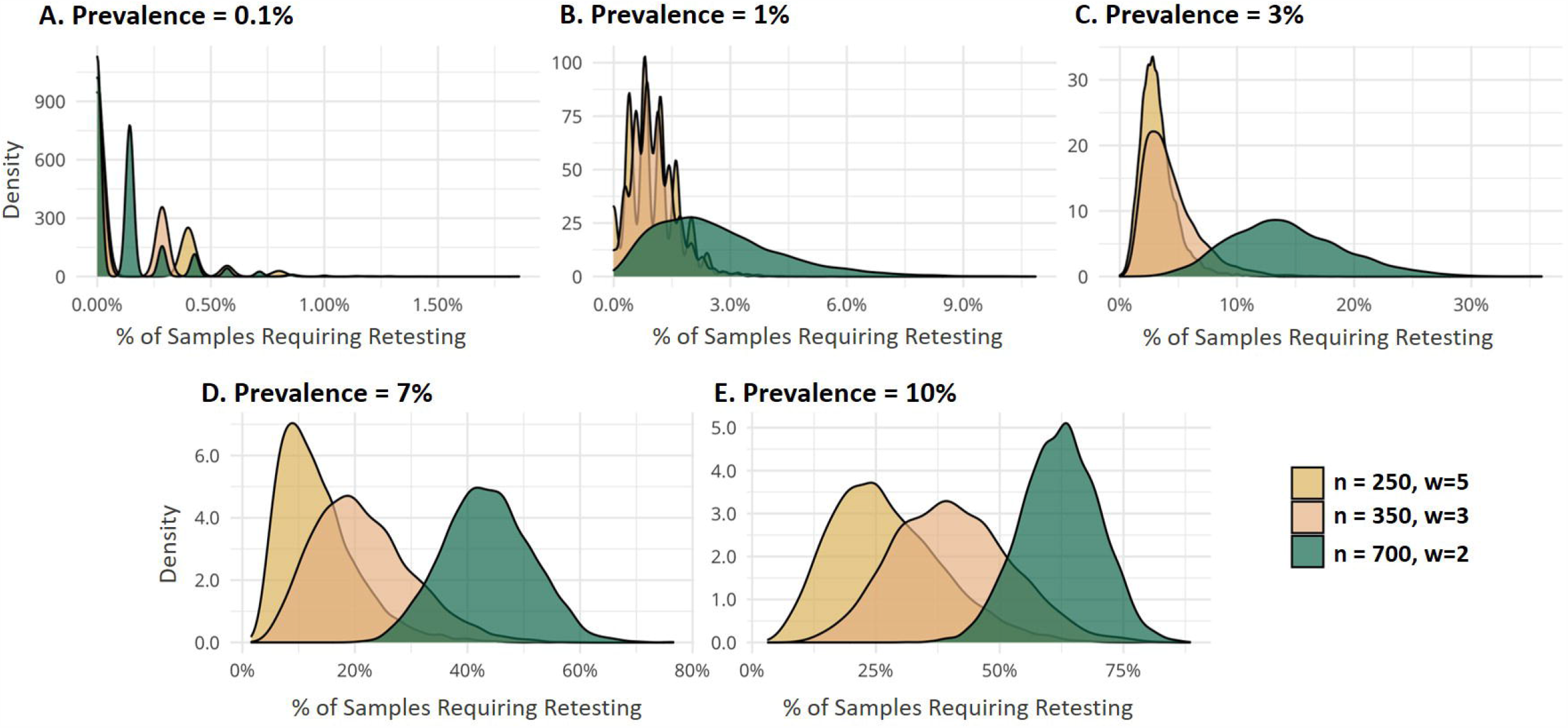
Proportion of Samples Requiring Retesting. A series of density plots demonstrating the number of retests required (i.e. where the matrix was unable to differentiate the true status of sample) after a single round of testing at each prevalence level. At very low prevalence levels (0.1- 1%) (plots A and B) there are only a small number of possible outcomes, therefore the density plots appear multi-modal.

The other models, n=350 and n=250, were notably more tolerant to an increase in population prevalence than the n=700 model. This is best demonstrated by the proportion of tests confirmed as negative (Figure 4). At a prevalence of 10% the proportion of true negatives that were confirmed in the n=700 matrix was just 38%. This is in comparison to the n=350 and n=250 models which were able to confirm an average of 60% and 73% as negative, respectively. The n=250 matrix was most tolerant to an increase in the population prevalence with most simulations proving relatively robust at prevalence of 7%, with an average of 87% of tests confirmed negative after a single run. At low population prevalence levels (0.1%-1%), all NAC matrices had a near perfect performance as measured by the proportion of true negatives identified after a single run (Figure 4).

**Figure 4.**
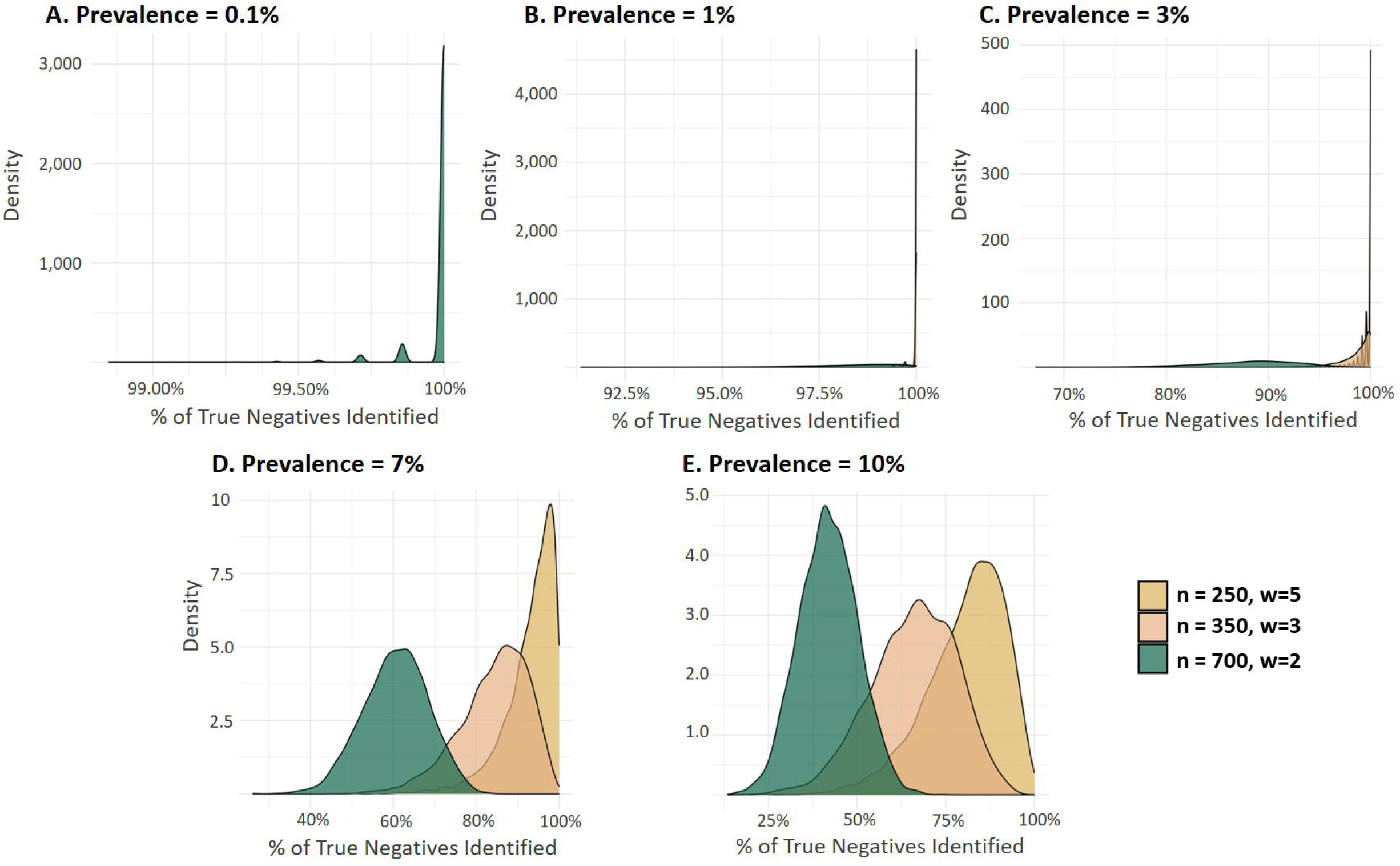
Proportion of True Negatives Identified. A series of Density plots demonstrating the proportion of samples confirmed negative as a percentage of the total number of true negatives.

The performance of each matrix was compared to a DS group testing strategy at the same population prevalence (Figure 5). Both the DS and NAC matrices saw their discriminatory ability deteriorate as the population prevalence increased. The n=250 and n=350 matrices were superior to DS approaches at all population prevalence levels except for when the prevalence reached 10% (Figure 5 and S1). At a population prevalence of 10% the DS approach was marginally superior for all matrices, although both group testing strategies at this prevalence level performed poorly, with over 350 retests required in both. The n=700 NAC matrix was the least robust, although it performed better than its DS counterpart at low prevalence levels (0.1-3%), its discriminatory ability was inferior once the population prevalence climbed above 7% (Figure 4).

**Figure 5.**
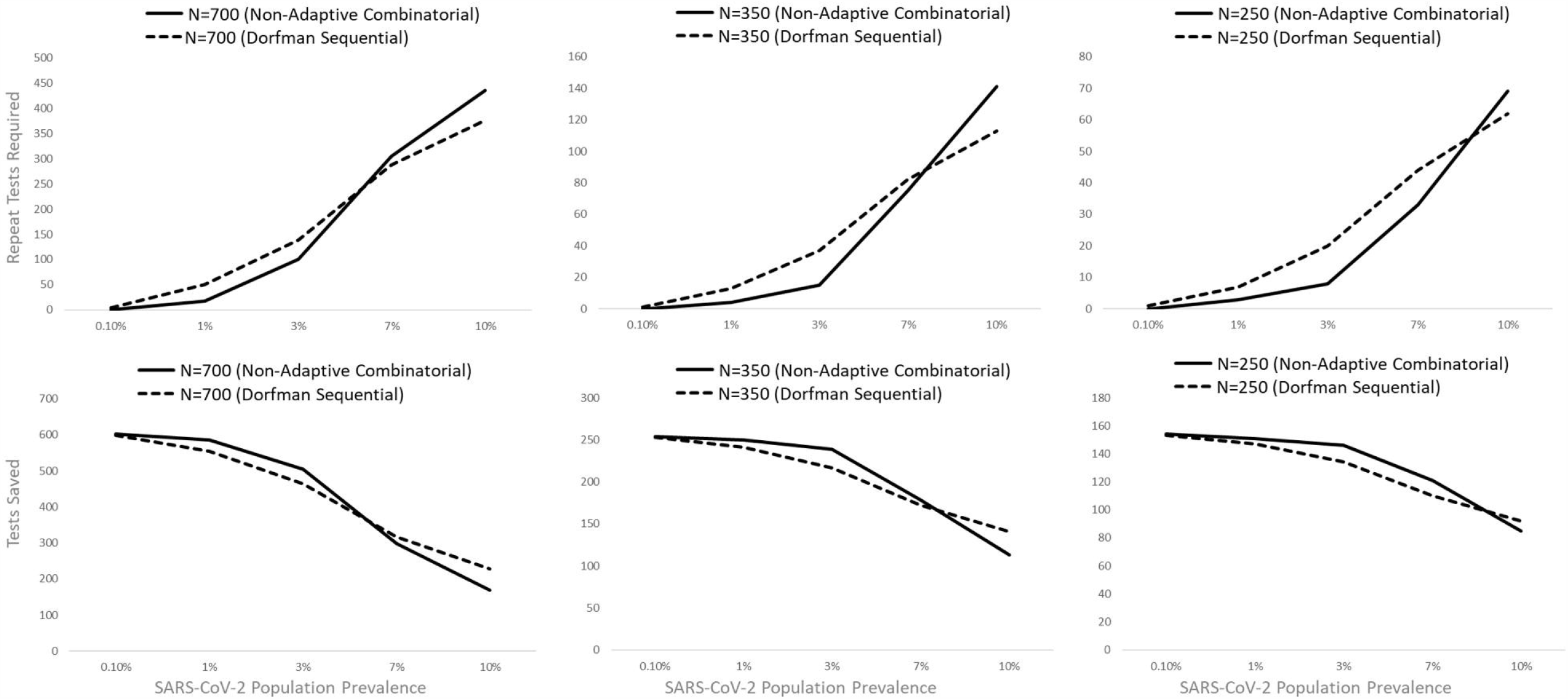
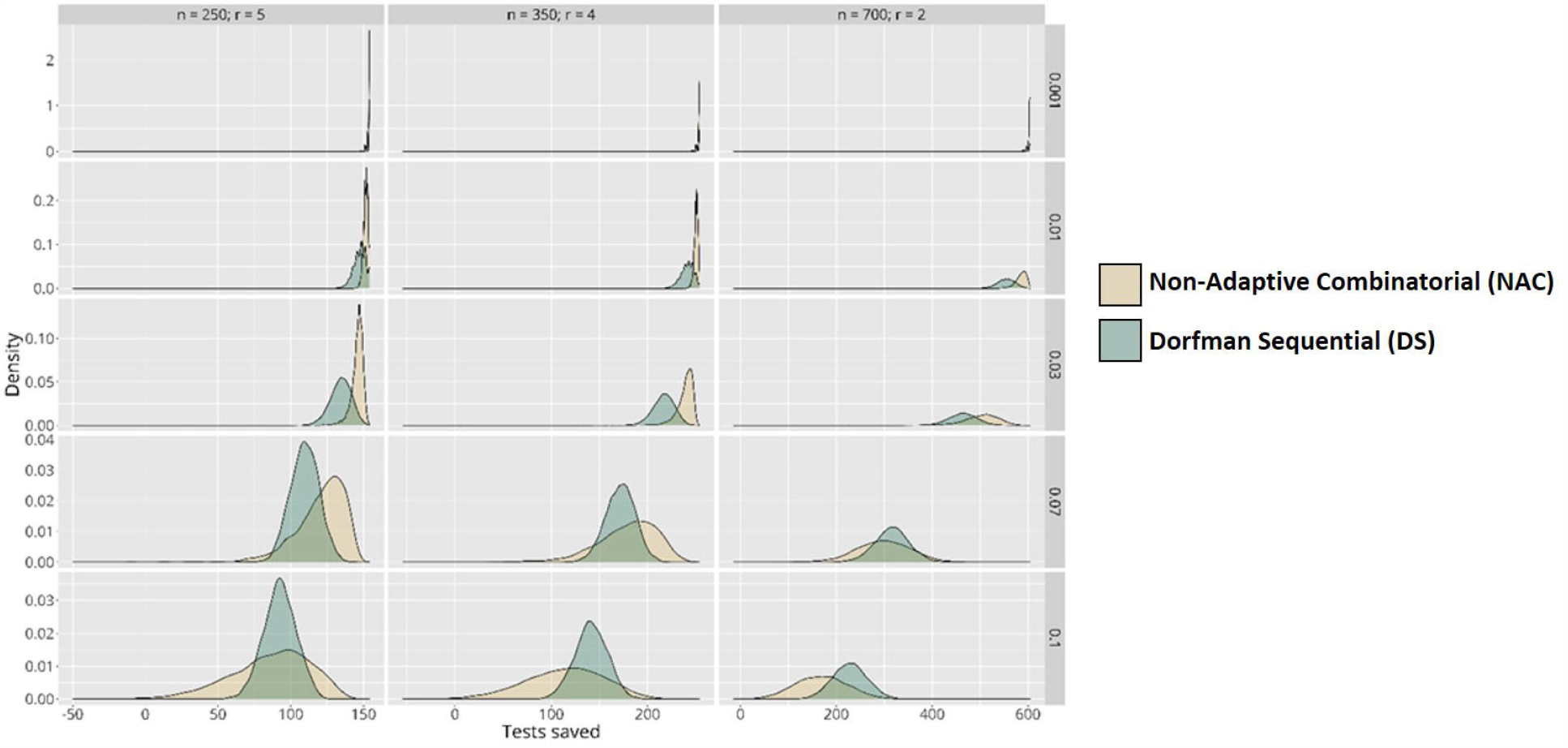
Non-adaptive Combinatorial Vs Dorfman Sequential Pooling Approaches. Matrices were tested at population prevalences of 0.1%, 1%, 3%, 7% and 10%. Performance statistics for the total number of tests saved (compared to monoplex approach) and the number of retests required are displayed.

## 4. Discussion

Since the SARS-CoV-2 outbreak began in late 2019, there have been over 13 million reported cases and few countries remain unaffected. Despite the global nature of the pandemic, the status of the outbreak differs markedly between nations and the prevalence in each nation is not homogenous. At the time of writing some countries, such as Brazil, find themselves part of an escalating outbreak while other nations, like the UK, are experiencing a steady decline in the number of cases each day. Given this variation, it is likely that the optimal testing strategy in one country will not be optimal in another country. Similarly, the most appropriate testing approach may change within a single country as the population prevalence changes or if there are certain settings, such as healthcare facilities, where the expected positivity rate is higher. A NAC group testing strategy offers a method that can be adapted based on the expected local or national population prevalence, increasing throughput, and saving reagents.

We designed three NAC matrices and tested their single round efficacy at varying population prevalences. When the prevalence was low, all the matrices performed well with only fractions of samples requiring re-testing. This is especially true in very low prevalence settings (0.1%) where, even in the least tolerant n=700 matrix, only one retest was required on average. However, as the population prevalence increased the performance of each matrix deteriorated. Even the most resilient matrix, n=250, was limited in its discriminatory ability when the population prevalence rose above 10%. Numerous reports early in the pandemic detailed programs designed to offer testing to symptomatic healthcare workers at hospitals across the UK.^13; 14^ In these trials, the positivity rate in symptomatic staff members ranged from 14-20%. Any pooling strategy, if the positivity rate were this high, would perform poorly and the number of re-tests required would become onerous, defeating the object of a group testing strategy. This is supported by the data presented here and we would therefore not recommend the use of pooling when the prevalence of SARS-CoV-2 is higher than 7%. Rather, the current monoplex approach would likely remain most practicable.

The data outlined in this work demonstrates that pooling becomes increasingly useful as the population prevalence of SARS-CoV-2 decreases. A recent trial at Barts Hospital, London, tested asymptomatic HCWs for 5 consecutive weeks with positivity rates ranging from 7.1% to 1.1%.^15^ The peak of 7.1% was recorded on the 30^th^ March 2020, one week after the UK-wide lockdown and at a time when community viral transmission was likely to have been at its highest. This changing prevalence demonstrates how a context specific adaptive testing strategy for staff testing might be deployed. Initially the most conservative matrix, n=250, should be used to establish the population prevalence. At a positivity rate of 7.1% this matrix would still be able to identify 90% of true negatives save approximately 121 tests relative to a monoplex approach, superior to a DS strategy at the same sample size. If the positivity rate were lower, this would then inform the choice of the matrix for subsequent testing rounds. As the rate falls, the utilization of less tolerant but higher throughput assays could be used, such as the n=700 matrix described here.

A maximum pool size of 15 was a formal condition for all the matrices outlined in this work. Based on previously published work, it is likely that this pool size could be increased without losing assay sensitivity. A recent correspondence outlined how pooling of up to 30 samples could be used to increase throughput without impacting diagnostic accuracy, although it was noted that borderline positive samples may escape detection in larger pools.^16^ We chose to test, and subsequently use, a pool size of 15 samples for two main reasons. First, this pool size will likely be more sensitive to borderline positive samples than larger pools, an important benefit when considering the importance of avoiding false negative results in HCWs who will be interacting with vulnerable patients. Secondly, although higher pool sizes increase the theoretical throughput of a single plate, the complexity of the assay design and processing also increases.

Critically, we propose these pooling approaches to improve throughput, save resource, and reduce the time that HCWs are waiting for their results. Typically, group testing strategies utilize subsequent rounds of group testing. These approaches are likely to be mathematically more efficient, as measured by the number of tests required to resolve all the samples, than our approach outlined here.^17^ However, more than one round of group testing introduces complexity, utilizing laboratory resources, and increases the time-to-result.

Any testing scheme will have positive and negative attributes which can be broadly split into throughput, reagent efficiency, speed, and complexity. The most suitable testing strategy will be context dependent. In some situations, such as widespread population screening, throughput will understandably be the dominant attribute and complex, multi-stage, centralized approaches can be used.^18^ We argue that in the context of HCW screening, a more subtle balance must be struck between speed and throughput, with complexity reduced if testing is being performed on a more local basis. We believe that the approach described here, an initial NAC screen followed by monoplex testing, provides a relatively high throughout system with good efficiency and the design can be varied depending on the local sample size and expected population prevalence.

We have created three distinct NAC matrices and tested their performance against other testing approaches. These matrices and the system to decode the results are freely accessible (www.samplepooling.com). At low to medium (0.1-7%) positivity rates, as would be expected in asymptomatic HCWs, the matrices are able to increase throughput and reduce the requirement for repeated testing compared to DS or standard monoplex schemes. The benefit of the approach outlined here is that laboratories can choose the matrix which most suits their current population prevalence and sample size, facilitating a context specific and data-driven testing approach.

## Data Availability

The matrices are freely available at www.samplepooling.com. The code underlying the decoding system and the matrix generator is publicly available.

https://github.com/duncstod/grouptesting

https://github.com/MCGM-Covid-19/matrix-decoder.github.io

